# COVID-19 vaccination and short-term mortality risk: a nationwide self-controlled case series study in The Netherlands

**DOI:** 10.1101/2024.12.11.24318790

**Authors:** Isabel AL Slurink, Annemarijn R de Boer, Marc JM Bonten, Miriam CJM Sturkenboom, PCJL Bruijning-Verhagen

## Abstract

Excess mortality during the COVID-19 pandemic partly exceeded COVID-19-related deaths, indicating that other causes may have contributed. We conducted a retrospective data-linkage study including all Dutch inhabitants to investigate the impact of COVID-19 vaccination on excess mortality using a modified self-controlled case series method. We found a 44% lower relative incidence of all-cause deaths in the first three weeks after the primary vaccination compared to more than three weeks after vaccination (IRR 0.56, 95%CI 0.54-0.57). This lower incidence was consistent across vaccine types, doses, genders, age groups, and individuals with or without prior SARS-CoV-2 infection or comorbidities, and for non-COVID-19 related deaths. For booster vaccinations, the relative incidence was similar (IRR 0.49, 95%CI 0.49-0.50). In comparison, we observed a 16-fold higher incidence of all-cause deaths in the three weeks following a registered positive SARS-CoV-2 infection compared to more than three weeks after infection (IRR 16.19, 95%CI 15.78-16.60). A lower relative incidence of short-term deaths following COVID-19 vaccination support that COVID-19 vaccination is not associated with the observed excess mortality.

## Introduction

The COVID-19 pandemic has significantly impacted global mortality rates, leading to an estimated 14.83 million excess deaths, which is 2.74 (2.44-3.06) times higher than the 5.42 million reported COVID-19 deaths ^1^. This excess mortality could be due to both the direct effects of SARS-CoV-2 infections, and indirect effects such as disruptions in healthcare systems and economic instability. In the Netherlands, three periods of excess mortality were observed in 2020 and 2021, each period having around 10% higher mortality than expected ^2^. The first two periods coincided with waves of COVID-19-reported deaths. However, during the third period of excess mortality in the fourth quarter of 2021, the excess mortality exceeded the COVID-19-reported deaths. This suggests that other causes of death may have contributed to the excess mortality. The difference was most pronounced for individuals aged 65 to 80.

COVID-19 vaccination has shown to be highly effective against symptomatic SARS-CoV2 infections in randomized clinical trials, as well as in observational studies showing reduced risk of severe illness and deaths from COVID-19 ^3-7^. In response to vaccination, the immune system is activated to generate an immune response. This temporary activation can, in rare cases, be accompanied by severe side effects. Several severe side effects of COVID-19 vaccination have been reported to occur in the weeks following vaccination. These include myocarditis and pericarditis, thrombosis and other rare cardiovascular events, as well as neurological complications ^8-12^. Whether COVID-19 vaccination contributed to excess mortality has been studied in several countries, consistently showing no increased risk for non-COVID-19 related mortality and a protective effect for COVID-19-related mortality ^5,6,13-15^. Nevertheless, these studies could not exclude the possibility of several forms of bias, which may occur due to prognostic differences between vaccinated and unvaccinated subjects ^2,15^.

In this context, the self-controlled case series (SCCS) design complements the current evidence by performing within-person comparisons, using each case (a person who died) as their own control. This automatically controls for time-fixed confounding, thus accounting for factors that vary by vaccination status such as frailty, comorbidities and socio-economic factors. An extension of the SCCS design was developed to quantify short-term mortality following COVID-19 vaccinations ^16^, which has been previously used in COVID-19 vaccine safety studies ^17-21^. This design focuses on the immediate period after vaccination, during which immune activation may increase the risk of severe side effects, resulting in death. Nevertheless, these studies consistently show a lower risk of death from any cause following vaccination compared to non-vaccination periods. Considering the protective effect of vaccination on COVID-19 mortality, assessment of non-COVID-19 mortality will further elucidate the causative role of COVID-19 vaccination in the observed excess deaths. Furthermore, the impact of previous SARS-CoV-2 infection, and presence of comorbidities, on these associations, remains understudied.

This study investigates the association between COVID-19 vaccine administration and (short-term) deaths from any cause using a SCCS approach, to assess if COVID-19 vaccination can explain the observed excess mortality during the COVID-19 pandemic. Furthermore, associations with non-COVID related deaths were assessed. Differences in type of COVID-19 vaccination (mRNA or non-mRNA), dose-effect and impact of sex, age, previous SARS-CoV-2 infection, and comorbidities were investigated. The latter is especially important due to potential lower vaccine effectiveness and higher risk of mortality among populations with comorbidities ^22^. The analysis was repeated for the associations between a SARS-CoV-2 infection and each of the outcomes.

## Methods

### Study design and population

We performed an retrospective study using a SCCS design and record linkage approach in the general Dutch population. The study included Dutch inhabitants registered in the General population Personal Records Database (BPR) governed by Statistics Netherlands (CBS). For the analysis on the effect of primary COVID-19 vaccination, all subjects who died between January 6, 2021, the first day of vaccine administration in the Netherlands, and November 18, 2022, the start of the booster campaign were included. For the analysis on the effect of booster vaccinations, we additionally included subjects who died until April 30, 2023, the last recorded date in the mortality database at time of the study. For the analysis on the effect of a positive registered SARS-CoV-2 infection, inhabitants who died between June 1, 2020 and December 31, 2021 were selected, during the period when tests were registered.

All data sources were linked by an unique pseudonymized identifier. CBS is bound by the European General Data Protection Regulation. In addition, CBS adheres to the privacy stipulations in the Statistics Netherlands Act, the European Statistics Code of Practice, and its own Code of conduct. Informed consent is not feasible for this study since all research data is pseudonymized. This study falls within the exceptions mentioned in section 5 of the code of conduct for medical research. This study was conducted according to ‘gedragscode gezondheidsonderzoek’, the principles of the Declaration of Helsinki (World Medical Association, 2013) and in accordance with the EU GDPR (General Data Protection Regulation). This study does not fall under the scope of the Dutch Medical Research Involving Human Subjects Act (WMO). It therefore does not require approval from an accredited medical ethics committee in the Netherlands. However, in the UMC Utrecht, an independent quality check has been carried out to ensure compliance with legislation and regulations (regarding Informed Consent procedure, data management, privacy aspects and legal aspects).

### Outcomes

The primary outcome of this study was death from any cause. As a secondary outcome, non-COVID-19 related death was examined. Notification of the cause, time and location of death is mandatory by Dutch law. Death certificates are processed and coded by CBS into The National Cause of Death Registry (1995-2023). From this registry, the date and cause of death were extracted. The event or disease that initiated the process of events that led to death is registered according to the International Statistical Classifications of Diseases and Related Health Problems, 10th revision (ICD-10) of the World Health Organization (WHO) ^23^. COVID-19 deaths were defined based on the ICD-10 codes for proven (U07.1) or suspected COVID-19 (not tested or virus not identified; U07.2). Non-COVID-19 related causes of death were grouped into six umbrella categories as used by Statistics Netherlands including neoplasms; circulatory system; respiratory system; mental, behavioral, and nervous system; external (non-natural) causes; and other natural causes ^2^.

### COVID-19 vaccination

COVID-19 vaccination data was obtained from the COVID-vaccinatie Informatie-en Monitoringssysteem (CIMS) (2020-2023) governed by the National Institute for Public Health and the Environment (RIVM). CIMS contains information on individuals who have consented to be registered in the vaccine registry. The COVID-19 vaccination campaign for the primary series started on 6 January 2021 in the Netherlands according to priority groups. The booster series started on 18 November 2022. Vaccination uptake was 87.4% among people aged 12 and over at the end of 2021 ^24^. Permission for registering vaccine administrations in the database was granted by 93% of participants for the primary series, 95% for the booster series, and 99% for the repeated dose. From this registry, the date of vaccination, vaccine batch, type of vaccination, vaccine dose, and administrator was extracted. The type of vaccine was categorized as mRNA (Pfizer/BioNTech, and Moderna) and non-mRNA or unknown (AstraZeneca, Janssen, Novavax, Valneva, Sanofi Pasteur, HIPRA, or vaccine type unknown).

### SARS-CoV-2 infection

Information about reported SARS-CoV-2 infections to the Dutch municipal health services were obtained from a database governed by the municipal health services [HPZone Lite van de gemeentelijke gezondheidsdiensten, GGDCOVID19BM] (2020-2021). At the time of the study-period, COVID-19 was a notifiable disease in the Netherlands meaning that all positive SARS-CoV-2 infections had to be reported to a municipal health service. From this registry, the registration date of a SARS-CoV-2 infection and type of SARS-CoV-2 test were extracted. A positive reporting within 90 days of a previous positive report was assumed to belong to the previous SARS-CoV-2 infection episode and excluded ^25^. At the start of 2021, the SARS-CoV-2 wildtype dominated in the Netherlands, the Alpha variant dominated in Spring 2021, and Delta dominated from June 2021 onwards until Omicron BA.1 replaced Delta at the end of the year. The primary series was considered completed and effective 14 days after a subject received two or three (only indicated for immunocompromised individuals) primary vaccinations with either Spikevax® (Moderna), Comirnaty® (Pfizer-BioNTech), Nuvaxovid® (Novavax), or Vaxzevria® (AstraZeneca), or as a single dose of the Janssen COVID-19 Vaccine, as per national guidelines at the time ^26^. Alternatively, a completed primary series could be defined as a single vaccine dose of the Moderna, Pfizer, Novavax or AstraZeneca vaccine in subjects with a documented prior SARS-Cov2 infection.

### Covariates

Sex and age group (10-20 years band) were extracted from the population registry governed by Statistics Netherlands. Comorbidities before the date of first vaccination or SARS-CoV-2 infection were defined on the basis of hospital admission information were extracted from the Dutch Hospital Discharge Registry (DHDR) governed by the Dutch Hospital Data (DHD) foundation. The hospital admission and discharge data, institutions number (pseudonymized), admission number (pseudonymized), and diagnoses recorded as ICD-10 code (as of 2014) and ICD-9 code (1995-2013) were extracted from this database ^23,27^. The comorbidities were specified according to the ECDC core protocol for COVID-19 vaccine effectiveness studies including asthma, immunodeficiency (including HIV infection) and organ transplant, cancer, mellitus, heart disease (excluding hypertension), hypertension, lung disease (excluding asthma), obesity, anemia, asplenia, chronic liver disease, neuromuscular disorders, renal diseases, dementia, stroke, rheumatologic diseases and tuberculosis ^28^.

### Statistical analysis

Data linkage and statistical analyses were conducted using R version 4.4.4.

A modified SCCS method was used to estimate the incidence rate ratio (IRR) and 95% confidence intervals (CI) of death in a predefined risk interval compared to a predefined control interval ^16^. This modification is designed to handle multiple event-dependent exposures, such as COVID-19 vaccine administrations, in relation to death as the event of interest. With death as the event of interest, the key assumptions of a standard SCCS are violated, as death precludes subsequent exposures and observation periods. This modified SCCS compares the risk of death during a predefined risk period following exposure to a reference period, defined as all observation time during which subsequent exposures could have occurred. This makes the end of the observation period independent of events. The model is estimated by iteratively reweighting observations to align with a counterfactual scenario in which no exposures can occur after death, ensuring that death does not censor exposures, as there are no exposures in this counterfactual scenario.

The SCCS models were fitted using a conditional Poisson regression model using a pseudo-likelihood method on a person-week level dataset ^16,29,30^. The IRRs for deaths in the risk interval relative to the control interval were estimated. The 95% CI for the parameters were obtained using the sandwich estimator in which the inverse Jacobian matrix surrounds the observed covariance matrix of the estimating functions ^31,32^. The length of each week in days was included as an offset in the model, as some weeks are not complete (such as if a vaccination occurs part way through a week). The risk interval was defined as day 1 to 21 days following COVID-19 vaccine administration, excluding day 0 from analyses, or as day 0 to 21 following a registered positive SARS-CoV-2 infection. We examined the first three weeks combined, and for each of the three weeks individually. The control interval was defined as all the following weeks to the end of the study. For vaccination, the risk weeks were further categorised by dose or considered for all doses combined. Due to the within-person comparison, time-invariant confounding, such as sociodemographic factors, comorbidities and health-seeking behaviour, is automatically controlled for. To account for time trends in mortality incidence due to seasonality and fluctuating SARS-Cov-2 infection rates, calendar time in two-week intervals was included as a covariate in the analysis. This time-trend adjustment is independent of vaccination status and thus included cases with no registered vaccine administration or infection to avoid bias ^16^.

### Subgroup analyses

Analyses were stratified by sex, age group at start of the observation period (12-29, 30-39, 40-49, 50-59, 60-69, 70-79, 80-89, ≥90) and presence of comorbidities (none, one, multiple). The subgroup aged <12 was removed from analyses to comply with privacy regulations of Statistics Netherlands. Analyses for the primary series were additionally stratified for vaccine type (mRNA or non-mRNA/unknown), and a prior registered positive SARS-CoV-2 infection before vaccination (yes/no). The analyses of each vaccine type included all individuals who received at least one dose of the vaccine type of interest. The doses were renumbered to correspond to the first, second, and third dose of the vaccine type of interest, while doses of other vaccine types were excluded by setting them as missing. Analyses for positive registered SARS-CoV-2 infection were additionally stratified by vaccination status at time of registration.

### Sensitivity analyses

Multiple sensitivity analyses were performed to assess robustness of findings or to test relevant secondary hypothesis. First, individuals with a positive SARS-CoV-2 infection in the 8 weeks before vaccination or during the exposure period were excluded. Second, calendar time adjustment was done using a restricted cubic spline to test whether this was a more realistic representation of confounding by calendar time effects. Third, we repeated the analysis without adjustment for calendar time, to evaluate its impact. Third, a 12-week risk periods was used to assess sensitivity to the specification of the risk period. Fourth, analyses were stratified according to administrator (municipal health services, general practitioner, or other) to account for possible difference in patient populations and to identify potential inconsistencies in data registration. Lastly, a, 2-, 4-, 7- and 14-day induction interval before the registration date of positive SARS-CoV-2 was applied (excluding observation time immediately preceding the index date) to account for potential delays in test registration.

## Results

A total of 493,382 deaths were registered among Dutch inhabitants of all ages between June 1, 2020 and December 31, 2023. The number of deaths each week by main cause of death is shown in **Figure 1**. COVID-19 related mortality was highest in January-February 2021, decreasing during the summer months, and surged again in November 2021. Mortality from other causes of deaths remained relatively stable throughout the study period. Trends in COVID-19 related mortality were more pronounced in older age groups and individuals with multiple comorbidities (**Supplemental figure 1A&C**).

**Figure 1.**
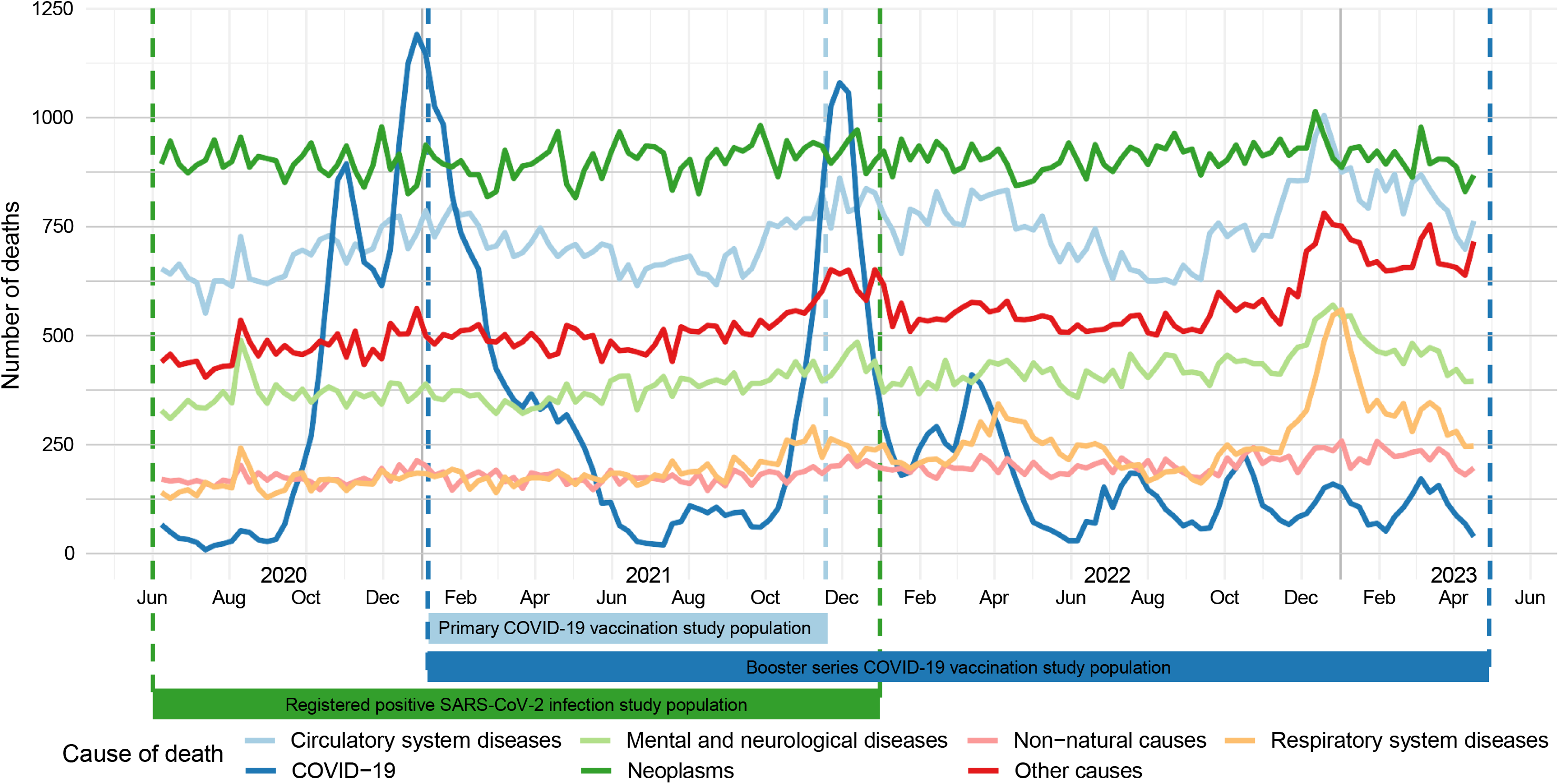
Number of deaths each week by main cause, among individuals who died between June 1, 2020 to April 30, 2023 (*n* = 493,382).

### Relative incidence of deaths following primary COVID-19 vaccination

A total of 75,421 individuals had received one or more doses of the primary COVID-19 vaccination and died between January 6, 2021, and November 18, 2022 (**Supplemental figure 2**). The mean age was 79.8 ± 11.8 years and 51.2% were female (**Table 1**). Most individuals received two doses (78.9%), the majority were mRNA vaccines (90.7%). More than half had more than one chronic condition (57.6%), with CVD, hypertension, cancer and lung disease being the most common. Characteristics according to the risk period after vaccination compared to individuals without registered vaccine administration are shown in **Supplemental table 1**. Differences between individuals who died within three weeks after vaccination and those who died in the period thereafter were minor. The distribution of primary vaccine administrations over time in the study population is shown in **Supplemental figure 3a**.

**Table 1.** Characteristics of individuals in the general Dutch population who received at least one COVID-19 vaccination and died from any cause between January 6, 2021 and November 18, 2021.

| Characteristic | Study Population ($n = 75,421$) |
| --- | --- |
| Mean age (SD), years | 79.8 (11.8) |
| Age group |  |
| <29 | 234 (0.3) |
| 30-39 | 277 (0.4) |
| 40-49 | 862 (1.1) |
| 50-59 | 3,145 (4.2) |
| 60-69 | 8,409 (11.1) |
| 70-79 | 18,469 (24.5) |
| 80-89 | 28,933 (38.4) |
| $\geq 90$ | 15,092 (20.0) |
| Women, n (%) | 38,612 (51.2) |
| Cause of death |  |
| Circulatory system diseases | 18,499 (24.5) |
| Neoplasms | 20,663 (27.4) |
| Respiratory system diseases | 4,983 (6.6) |
| Mental and neurological diseases | 11,400 (15.1) |
| Non-natural causes | 4,322 (5.7) |
| Other causes | 12,168 (16.1) |
| COVID-19 | 3,386 (4.5) |
| Number of vaccine administrations primary series |  |
| One dose | 15,712 (20.8) |
| Two doses | 59,492 (78.9) |
| Three doses<sup>1</sup> | 217 (0.3) |
| Vaccine type, first vaccine |  |
| mRNA | 68,373 (90.7) |
| non-mRNA or unknown | 7,045 (9.3) |
| Vaccine type, second vaccine<sup>2</sup> |  |
| mRNA | 55,543 (73.6) |
| non-mRNA or unknown | 4,168 (5.5) |
| Vaccine type, third vaccine<sup>1</sup> |  |
| mRNA | 164 (0.2) |
| non-mRNA or unknown<sup>2</sup> | 55 (0.1) |
| Risk period after vaccination |  |
| $\leq 3$ weeks | 8,073 (10.7) |
| $> 4$ weeks | 67,348 (89.3) |
| Presence of comorbidities<sup>3</sup> |  |
| No | 17,176 (22.8) |
| One | 14,795 (19.6) |
| Multiple | 43,450 (57.6) |
| Number of registered positive SARS-CoV-2 infections |  |
| 0 | 68,000 (90.2) |
| 1 | 7,344 (9.7) |
| 2 or 3 | 77 (0.1) |

<sup>1</sup> Indicated for immunocompromised individuals
<sup>2</sup> Rounded to nearest fifth due to low counts.
<sup>3</sup> Number and frequencies of specific comorbidities (e.g. CVD, hypertension, cancer, lung disease) shown in Supplemental table 1.

The relative incidence of all-cause deaths in the three-weeks following any dose of the primary vaccination, compared with the period thereafter, was 0.56 (95%CI 0.54-0.57) (**Table 2**). The relative incidence varied within the risk interval from 0.33 (95%CI 0.31-0.34) in week one, to 0.56 (95%CI 0.54-0.58) in week two, to 0.73 (95%CI 0.70-0.75) in week three. The relative incidence was comparable across different vaccine doses of the primary series (range 0.55–0.61) and across sexes (0.55–0.56), age groups (0.48–0.64), presence of comorbidities (0.51–0.57), and prior COVID-19 infection (0.56–0.61). A slightly lower relative incidence was found for non-mRNA or unknown vaccine types (IRR 0.40, 95%CI 0.37-0.43) compared to mRNA vaccine types (IRR 0.53, 95%CI 0.51-0.54). This was only evident for the first dose (IRR 0.34, 95%CI 0.31-0.37) and not for the second and third dose (IRR 0.54, 95%CI 0.48-0.60 and IRR 0.69, 95%CI 0.24-1.97, respectively). Associations were similar considering non-COVID-19 related deaths as outcome.

**Table 2.** Relative incidence of all-cause and non-COVID-19 related deaths in the 3-week risk period following primary series vaccination, by risk period and subgroup.

|  | All-cause deaths | Non-COVID-19 related deaths |
| --- | --- | --- |
|  | Relative incidence<br>(95% CI) | Relative incidence<br>(95% CI) |
| **All doses combined** |  |  |
| Weeks 1-3 | 0.56 (0.54-0.57) | 0.55 (0.53-0.56) |
| Week 1 | 0.33 (0.31-0.34) | 0.34 (0.33-0.36) |
| Week 2 | 0.56 (0.54-0.58) | 0.55 (0.53-0.57) |
| Week 3 | 0.73 (0.70-0.75) | 0.69 (0.66-0.71) |
| **First dose** |  |  |
| Weeks 1-3 | 0.56 (0.55-0.58) | 0.54 (0.52-0.55) |
| Week 1 | 0.31 (0.29-0.34) | 0.34 (0.31-0.36) |
| Week 2 | 0.55 (0.53-0.58) | 0.53 (0.50-0.56) |
| Week 3 | 0.76 (0.73-0.79) | 0.68 (0.65-0.72) |
| **Second dose** |  |  |
| Weeks 1-3 | 0.55 (0.53-0.57) | 0.56 (0.54-0.58) |
| Week 1 | 0.34 (0.32-0.37) | 0.36 (0.33-0.39) |
| Week 2 | 0.57 (0.54-0.60) | 0.58 (0.55-0.61) |
| Week 3 | 0.69 (0.65-0.72) | 0.70 (0.66-0.73) |
| **Third dose** |  |  |
| Weeks 1-3 | 0.61 (0.44-0.84) | 0.56 (0.40-0.79) |
| Week 1 | 0.18 (0.04-0.76) | 0.18 (0.04-0.78) |
| Week 2 | 0.23 (0.13-0.42) | 0.24 (0.13-0.44) |
| Week 3 | 0.60 (0.40-0.91) | 0.57 (0.37-0.88) |
| **Vaccine type, mRNA** |  |  |
| All doses combined | 0.53 (0.51-0.54) | 0.52 (0.50-0.53) |
| First dose | 0.54 (0.52-0.55) | 0.51 (0.49-0.52) |
| Second dose | 0.52 (0.50-0.54) | 0.53 (0.51-0.55) |
| Third dose | 0.57 (0.40-0.82) | 0.50 (0.34-0.74) |
| **Vaccine type, non-mRNA or unknown** |  |  |
| All doses combined | 0.40 (0.37-0.43) | 0.41 (0.38-0.44) |
| First dose | 0.34 (0.31-0.37) | 0.35 (0.31-0.38) |
| Second dose | 0.54 (0.48-0.60) | 0.53 (0.48-0.59) |
| Third dose | 0.69 (0.24-1.97) | 0.66 (0.23-1.89) |
| **Subgroup analyses, all doses combined** |  |  |
| **Sex** |  |  |
| Men | 0.56 (0.54-0.58) | 0.55 (0.53-0.57) |
| Women | 0.55 (0.53-0.57) | 0.54 (0.52-0.56) |
| **Age group** |  |  |
| 12-29 | 0.54 (0.37-0.80) | 0.53 (0.36-0.78) |
| 30-39 | 0.64 (0.46-0.90) | 0.64 (0.46-0.90) |
| 40-49 | 0.56 (0.45-0.68) | 0.55 (0.45-0.67) |
| 50-59 | 0.48 (0.42-0.54) | 0.48 (0.42-0.54) |
| 60-69 | 0.53 (0.50-0.57) | 0.54 (0.50-0.58) |
| 70-79 | 0.54 (0.52-0.57) | 0.53 (0.50-0.56) |
| 80-89 | 0.53 (0.51-0.56) | 0.53 (0.51-0.56) |
| ≥90 | 0.51 (0.48-0.54) | 0.53 (0.50-0.56) |
| **Presence of chronic disease** |  |  |
| None | 0.57 (0.54-0.60) | 0.55 (0.52-0.58) |
| One | 0.51 (0.48-0.54) | 0.50 (0.47-0.53) |
| Multiple | 0.57 (0.55-0.59) | 0.56 (0.54-0.58) |
| **Prior COVID-19 infection** |  |  |
| Yes | 0.61 (0.55-0.67) | 0.59 (0.53-0.66) |
| No | 0.56 (0.54-0.57) | 0.55 (0.53-0.56) |

### Relative incidence of deaths following booster series COVID-19 vaccination

Between January 6, 2021, and April 30, 2023, a total of 396,765 deaths were registered, linked to 299,935 records of individuals who received at least one vaccine (**Supplemental figure 4**; **Supplemental table 2)**. The first booster dose was administered starting November 2021, the second starting January 2022, the third in April and the fourth started in September 2022 (**Supplemental figure 3b**). Consistent with main analysis, the relative incidence of all-cause deaths was significantly lower for all doses combined (IRR 0.49, 95%CI 0.49-0.50) and stratified per primary or booster dose (IRR ranging from 0.46-0.51) (**Table 3**). The relative incidence was similar across men and women, and for age groups, and was consistent for non-COVID-19 related deaths.

**Table 3.**
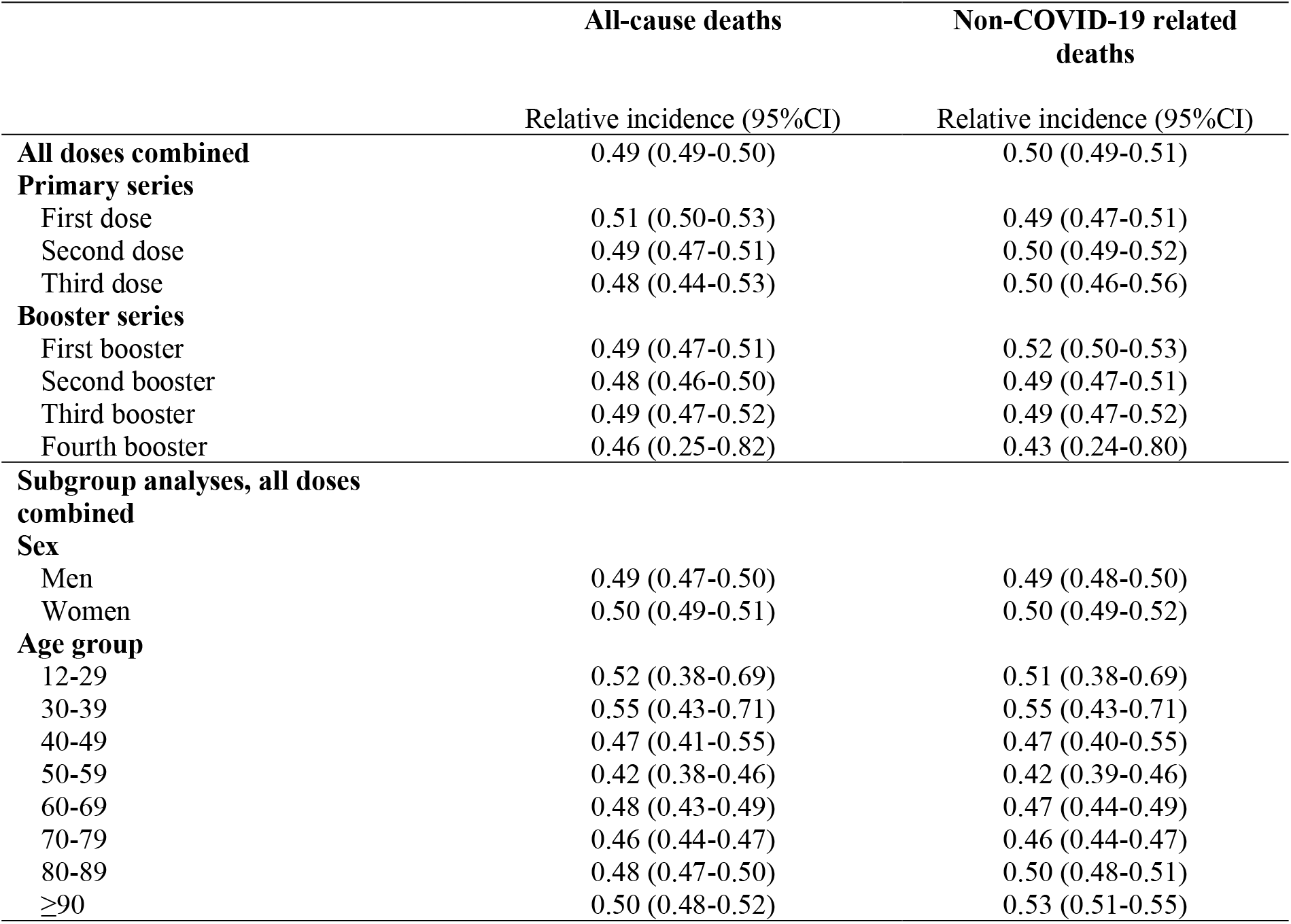
Relative incidence of all-cause deaths in the 3-week risk period following primary and booster series vaccination.

|  | All-cause deaths | Non-COVID-19 related deaths |
| --- | --- | --- |
|  | Relative incidence (95%CI) | Relative incidence (95%CI) |
| **All doses combined** | 0.49 (0.49-0.50) | 0.50 (0.49-0.51) |
| **Primary series** |  |  |
| First dose | 0.51 (0.50-0.53) | 0.49 (0.47-0.51) |
| Second dose | 0.49 (0.47-0.51) | 0.50 (0.49-0.52) |
| Third dose | 0.48 (0.44-0.53) | 0.50 (0.46-0.56) |
| **Booster series** |  |  |
| First booster | 0.49 (0.47-0.51) | 0.52 (0.50-0.53) |
| Second booster | 0.48 (0.46-0.50) | 0.49 (0.47-0.51) |
| Third booster | 0.49 (0.47-0.52) | 0.49 (0.47-0.52) |
| Fourth booster | 0.46 (0.25-0.82) | 0.43 (0.24-0.80) |
| **Subgroup analyses, all doses combined** |  |  |
| **Sex** |  |  |
| Men | 0.49 (0.47-0.50) | 0.49 (0.48-0.50) |
| Women | 0.50 (0.49-0.51) | 0.50 (0.49-0.52) |
| **Age group** |  |  |
| 12-29 | 0.52 (0.38-0.69) | 0.51 (0.38-0.69) |
| 30-39 | 0.55 (0.43-0.71) | 0.55 (0.43-0.71) |
| 40-49 | 0.47 (0.41-0.55) | 0.47 (0.40-0.55) |
| 50-59 | 0.42 (0.38-0.46) | 0.42 (0.39-0.46) |
| 60-69 | 0.48 (0.43-0.49) | 0.47 (0.44-0.49) |
| 70-79 | 0.46 (0.44-0.47) | 0.46 (0.44-0.47) |
| 80-89 | 0.48 (0.47-0.50) | 0.50 (0.48-0.51) |
| ≥90 | 0.50 (0.48-0.52) | 0.53 (0.51-0.55) |

### Relative incidence of deaths following a registered positive SARS-CoV-2 infection

Between June 1, 2020, and December 31, 2021, a total of 263,518 deaths were registered which were linked to 34,066 registered positive SARS-CoV-2 infections among 33,875 unique individuals (**Supplemental figure 5**). For 36.6% of these individuals, a vaccine administration was registered, and COVID-19 was the main cause of death (65.9%) (**Table 4**). More men, at higher age and with more comorbidities died within three weeks after infection, compared to those who died in the period thereafter or had no infection registered (**Supplemental table 3**). The numbers of registered infections over time in the study population are shown in **Supplemental figure 6**. There were peaks in infections in November 2020, January 2021 and in November-December 2021. Most individuals had one registered infection.

**Table 4.** Characteristics of individuals in the general Dutch population with a registered positive SARS-CoV-2 infection and who died from any cause between June 1, 2020 and December 31, 2021.

| Characteristic | Study population (n = 33,875) |
| --- | --- |
| Mean age (SD), years | 79.6 (11.7) |
| Age group, n (%) |  |
| <29 | 149 (0.4) |
| 30-39 | 120 (0.4) |
| 40-49 | 377 (1.1) |
| 50-59 | 1,334 (3.9) |
| 60-69 | 3,578 (10.6) |
| 70-79 | 8,836 (26.1) |
| 80-89 | 13,331 (39.4) |
| >=90 | 6,150 (18.2) |
| Women, n (%) | 16,146 (47.7) |
| Cause of death |  |
| Circulatory system diseases | 2,819 (8.3) |
| Neoplasms | 2,664 (7.9) |
| Respiratory system diseases | 669 (2.0) |
| Mental and neurological diseases | 1,917 (5.7) |
| Non-natural causes | 957 (2.8) |
| Other causes | 2,513 (7.4) |
| COVID-19 | 22,336 (65.9) |
| Number of registered COVID-19 infections |  |
| 1 | 33,685 (99.4) |
| 2 or 3 | 190 (0.6) |
| Type of COVID-19 test, first test<sup>1,2</sup> |  |
| PCR | 33,771 (99.8) |
| IgG | 21 (0.1) |
| Self-test | 43 (0.1) |
| Risk period after infection |  |
| ≤3 weeks | 21,243 (62.7) |
| >4 weeks | 12,632 (37.3) |
| Presence of comorbidities<sup>3</sup> |  |
| No | 10,215 (30.2) |
| One | 6,694 (19.8) |
| Multiple | 16,966 (50.1) |
| Vaccin type, first vaccine |  |
| mRNA | 11,607 (34.3) |
| non-mRNA or unknown | 787 (2.3) |
| Vaccin type, second vaccine |  |
| mRNA | 8,844 (26.1) |
| non-mRNA or unknown | 484 (1.4) |
| Vaccin type, third vaccine |  |
| mRNA | 194 (0.6) |
| non-mRNA or unknown<sup>4</sup> | 10 (0.0) |

<sup>1</sup> Antigen and IgM tests not shown due to low counts.
<sup>2</sup> All second and third tests were PCR.
<sup>3</sup> Number and frequencies of specific comorbidities (e.g. CVD, hypertension, cancer, lung disease) shown in Supplemental table 3.
<sup>4</sup> Rounded to nearest fifth due to low counts.

A higher relative incidence of all-cause deaths was observed in the three-weeks after a registered positive SARS-CoV-2 infection, compared to the period thereafter (IRR 16.19, 95%CI 15.78-16.60) (**Table 5**). The relative incidence was highest in the second week (IRR 22.63, 95%CI 21.97-23.31) and decreased in the third week (IRR 9.92, 95%CI 9.57-10.28). The relative incidence was lower for individuals with a registered vaccination within 6 months before infection (IRR 6.09, 95%CI 5.47-6.78) than individuals without registered vaccination or a vaccination longer than 6 months before infection (16.90, 95%CI 16.47-17.33). The relative incidence increased with age, ranging from 2.69 (95%CI 1.74-4.16) in those aged 12-29, to 20.60 (95%CI 19.77-21.46) in individuals aged 80-89. Furthermore, the relative incidence was slightly higher in men (IRR 18.00, 95%CI 17.37-18.65) compared to women (IRR 14.43, 95%CI 13.92-14.96), and in individuals with multiple comorbidities (IRR 17.34, 95%CI 16.73-17.97) compared to one (IRR 14.29, 95%CI 13.52-15.11) or none (IRR 15.26, 95%CI 14.57-15.98). The associations were substantially weaker when non-COVID-19 related deaths were used as the outcome.

**Table 5.** Relative incidence of all-cause deaths in the 3-week risk period following a registered positive SARS-CoV-2 infection, by risk period, vaccination status, and subgroups.

|  | **All-cause deaths** | **Non-COVID-19 related deaths** |
| --- | --- | --- |
|  | Relative incidence (95%CI) | Relative incidence (95%CI) |
| **All doses combined** |  |  |
| Weeks 1-3 | 16.19 (15.78-16.60) | 2.14 (2.03-2.26) |
| Week 1 | 16.99 (16.46-17.54) | 2.38 (2.19-2.57) |
| Week 2 | 22.63 (21.97-23.31) | 2.31 (2.14-2.51) |
| Week 3 | 9.92 (9.57-10.28) | 1.78 (1.64-1.94) |
| **Registered COVID-19 vaccination within 6 months before time of infection** |  |  |
| Weeks 1-3 | 6.09 (5.47-6.78) | 1.63 (1.35-1.97) |
| Week 1 | 6.75 (5.95-7.67) | 1.84 (1.43-2.37) |
| Week 2 | 7.99 (7.06-9.04) | 1.72 (1.32-2.24) |
| Week 3 | 3.89 (3.38-4.48) | 1.37 (1.05-1.79) |
| **No registered COVID-19 vaccination or vaccinated over 6 months prior to time of infection** |  |  |
| Weeks 1-3 | 16.90 (16.47-17.33) | 2.19 (2.07-2.32) |
| Week 1 | 17.63 (17.07-18.21) | 2.42 (2.23-2.64) |
| Week 2 | 23.73 (23.03-24.46) | 2.38 (2.18-2.59) |
| Week 3 | 10.33 (9.96-10.72) | 1.83 (1.67-1.99) |
| **Sex** |  |  |
| Men | 18.00 (17.37-18.65) | 2.57 (2.38-2.78) |
| Women | 14.43 (13.92-14.96) | 1.80 (1.66-1.94) |
| **Age group** |  |  |
| 12-29 | 2.69 (1.74-4.16) | 1.28 (0.71-2.31) |
| 30-39 | 5.30 (3.45-8.12) | 2.19 (1.13-4.23) |
| 40-49 | 5.59 (4.37-7.14) | 1.83 (1.23-2.72) |
| 50-59 | 6.43 (5.64-7.32) | 1.77 (1.37-2.28) |
| 60-69 | 9.50 (8.78-10.28) | 2.12 (1.80-2.50) |
| 70-79 | 17.66 (16.79-18.56) | 2.58 (2.31-2.88) |
| 80-89 | 20.60 (19.77-21.46) | 2.15 (1.96-2.35) |
| ≥90 | 15.68 (14.79-16.63) | 1.82 (1.60-2.06) |
| **Presence of chronic disease** |  |  |
| None | 15.26 (14.57-15.98) | 2.05 (1.84-2.28) |
| One | 14.29 (13.52-15.11) | 2.16 (1.92-2.42) |
| Multiple | 17.34 (16.73-17.97) | 2.14 (1.98-2.31) |
| **Excess mortality wave<sup>1</sup>** |  |  |
| Wave 2 | 8.30 (7.81-8.81) | 1.83 (1.61-2.07) |
| Wave 3 | 6.59 (6.14-7.08) | 1.77 (1.55-2.02) |

<sup>1</sup> Study period defined according to waves of excess mortality during the COVID-19 pandemic in the Netherlands <sup>2</sup>. Only registered positive SARS-CoV-2 infections and deaths recorded occurring during these periods were included in the analyses. The first wave of excess mortality, from 23-03-2020 to 27-04-2020, occurred before the start of SARS-CoV-2 registration. The second wave from 21-09-2020 to 18-01-2021, when a total of 60,644 individuals died. The third wave was from 16-08-2021 to 27-12-2021, when a total of 65,914 individuals died.

### Sensitivity analyses

The results were robust in sensitivity analysis without adjustment for calendar time, when using a restricted cubic spline for calendar time adjustment, and when excluding individuals who had a registered positive SARS-CoV-2 infection in the 8 weeks before vaccination or during the exposure period (**Supplemental table 4**). The relative incidence was slightly stronger for individuals vaccinated by municipal health services (IRR 0.46, 95%CI 0.44-0.48) or general practitioners (IRR 0.55, 95%CI 0.52-0.59), compared to those vaccinated by other administrators (IRR 0.77, 95%CI 0.74-0.80). The relative incidence was slightly reduced to 0.71 (95%CI 0.70-0.72) when considering a twelve-week risk period for all vaccine doses combined, compared to 0.56 (95%CI 0.54-0.57) in the three-week risk period analysed in the main analysis. Similarly, the relative incidence in the twelve-week risk period after a registered positive SARS-CoV-2 infection was reduced compared to main analysis (8.28, 95%CI 8.05-8.51 versus 16.19, 95%CI 15.78-16.60). Furthermore, setting the induction interval to 2, 4, 7 or 14 days before the registration date attenuated the relative incidence (IRR 14.79, 13.02, 9.91 and 2.54, respectively).

## Discussion

In this registry-based, nation-wide study in the Netherlands, a 44% lower incidence of all-cause and non-COVID-19 related deaths was observed in the first three weeks after any dose of the primary series vaccine administration compared to subsequent periods. This reduced incidence of all-cause deaths was most pronounced in the first week after vaccine administration and was slightly greater for non-mRNA or unknown vaccine types compared to mRNA vaccine types. The reduction was consistent across different vaccine doses, by sex, age groups, presence of comorbidities and registered prior COVID-19 infection. A lower relative incidence of deaths was also found following booster doses, consistent across different doses, as well as by sex and age groups. By contrast, we observed a 16-fold higher incidence of all-cause deaths after a registered positive SARS-CoV-2 infection, most notable in older age groups, and slightly higher in men and those with multiple comorbidities. The relative incidence ratios were slightly attenuated with a longer 12-week risk period.

### Results in context

Our results align with other SCCS studies conducted between 2020 and 2022, all reporting lower relative incidences of deaths shortly after vaccination compared with subsequent periods, consistent across demographic groups, vaccine doses, and types ^17-21^. A US cohort study (*n* = 18,000) found a substantial lower relative incidence of all-cause and non-COVID-19 deaths in the 14-day or 28-day risk interval following first or second vaccination (IRR range 0.23-0.55) ^18^. In a nation-wide registry in Italy (*n* = 2,167), the relative incidence of all-cause deaths within 30-days following vaccination was 0.76 (95%CI 0.70-0.83) ^19^. Using registry data in an urban city of Japan (*n* = 126), mRNA vaccination was associated with lower relative incidence of all-cause deaths within 3 weeks following the first and second dose (IRR 0.38, 95%CI 0.29-0.49 and 0.43, 032-0.56, respectively) ^20^. In a nationwide registry study in adult Singaporeans (*n* = 3,137,210), the relative incidence of all-cause deaths 31-days after any mRNA booster dose was 0.44 (0.40–0.48) ^17^. Furthermore, in a registry-study among 12 to 29 year-old individuals in England (*n* = 3,807), death rates were lower in the twelve weeks post-vaccination period for three doses combined (IRR 0.88, 95%CI 0.80-0.97) ^21^, with stronger associations for the second and third doses compared to the first, and for males compared to females.

Jointly, the evidence consistently points towards reduced mortality in the weeks following vaccination. This is a reassuring finding in the context of the observed and still partly unexplained excess mortality during the COVID-19 pandemic. If vaccination was causally related to an increase in mortality, the risk of death would be expected to peak shortly after vaccination and decline thereafter. In our study we observed the opposite; the risk of death was lowest shortly after vaccination. The lower death rate shortly after vaccination can be partly attributed to a time-varying ‘healthy vaccinee effect’ ^33^, such as the tendency for vaccinations to be administered during periods without illness, especially as fever is a contra-indication for vaccination ^34^. This unmeasured time-varying confounding is not fully addressed in SCCS designs, which also explains the similar low relative incidence rates for non-COVID-19 deaths. This healthy vaccinee effect may overestimate the reduction in deaths after vaccination to some extent and point estimates should therefore be interpreted with some caution.

We observed a slightly more pronounced incidence rate reduction for the first dose of non-mRNA or unknown vaccine types compared to mRNA vaccines, although this comparison is limited as only few people received these vector vaccines. Vector vaccines are no longer administered in the Netherlands due to high effectiveness of mRNA vaccines with lower incidence of adverse events, and logistic considerations. Prior studies have not demonstrated differences in all-cause ^19,21^, or excess mortality^35^, by vaccine type.

Our findings of high relative incidence of deaths following SARS-CoV-2 infection are also consistent with SCCS studies from Singapore and England ^17,21^. In our study, the relative incidence of deaths was highest in the second week after infection, in line with the typical clinical course of severe COVID-19 caused by SARS-CoV-2 wildtype, Alpha and Delta variants in the initial waves of the pandemic ^36,37^. This course typically begins with respiratory symptom onset in the initial phase, followed by a dramatic worsening due to a cytokine storm, leading to hospitalization and death within one to two weeks from symptom onset. The relative incidence of deaths following a positive SARS-CoV-2 infection was substantially lower among vaccinated individuals compared to unvaccinated individuals. A still somewhat higher incidence of deaths after a positive registered infection compared to control weeks in vaccinated individuals may reflect incomplete vaccine effectiveness and potential waning immunity from the primary series, particularly in older adults or those with pre-existing conditions ^38^, as well as selection bias, as testing might have been associated with higher severity of infections. Nevertheless, only a slight proportion of all infections was among vaccinated individuals, mostly registered in November 2021, coinciding with a peak in all-cause mortality.

### Strength and limitations

Strengths of our study include the nation-wide coverage of vaccination and mortality records, inclusion of data on previous SARS-CoV-2 infections and chronic medical conditions across all age groups, providing considerable sample size and generalizable findings.

The results should be interpreted considering the following limitations. First, while the date of death registration in the Netherlands is highly accurate, there is uncertainty in the reliability of the recorded primary cause of death ^39^. In a proportion of non-COVID-19 related deaths, SARS-CoV-2 infection may have had an unrecognized role in the causal pathway to death. Misclassification of non-COVID-19 related deaths could occur in both directions, potentially attenuating the observed associations. Despite, the associations with non-COVID-19 related death as the outcome remained substantial, suggesting that this misclassification is unlikely to have significantly altered our findings. Second, the national vaccination database is not complete as 7% of the primary series and 5% of the booster recipients did not give consent for registration. As a result, some cases without registered vaccine administration were, in fact, vaccinated. While this may have resulted in exclusion of some vaccinated subjects in our case series, this misclassification does not influence the IRR estimates, as cases without registered vaccine administrations are not part of the analysis. Routine procedure was to ask for consent for registration before the vaccine was administered, and therefore the probability of non-registration is unrelated to the likelihood of death following vaccination. Nevertheless, the associations between SARS-CoV-2 infection and mortality in individuals without a registered vaccine administrations are likely to be biased towards the null ^15,35^. Third, the median duration between the first and second primary vaccine was 5 weeks (IQR 4 - 5.1 weeks), limiting the longer-term risk assessment following the first dose. The duration of the risk interval affects the associations, as shown in sensitivity analyses with a 12-week risk interval. Lastly, routine testing for SARS-CoV-2 upon hospital admission could have inflated the associations, particularly during the first week after infection. The exact onset date of the infection is often unclear, complicating correct specification of the risk period. However, the results were robust in sensitivity analyses including induction days varying between 2 and 14 days.

## Conclusion

Taken together, our results provide additional evidence for absence of increased risk of death after COVID-19 vaccination, thereby excluding vaccination as a cause of observed excess mortality between January 2021 up to April 2023. The results indicate that for both all-cause and non-COVID-19-related deaths, the relative incidence in the three weeks following vaccination is lower compared to the period more than three weeks after vaccination. The reduced risk of death remained evident for up to 12 weeks post-vaccination. In comparison, the relative incidence of all-cause death in the three weeks following a SARS-CoV-2 infection was higher compared to the period more than three weeks after infection. Our findings can inform communication strategies about safety of COVID-19 vaccination, emphasizing consistency across demographic groups, vaccine types, and doses.

## Data Availability

All data are available within CBS Microdata and can be made available under strict conditions: Microdata: Conducting your own research https://www.cbs.nl/en-gb/our-services/customised-services-microdata/microdata-conducting-your-own-research#:~:text=Microdata%20are%20linkable%20data%20at,strict%20conditions%20for%20statistical%20research

https://github.com/isabelslurink/SCCS_COVID19_mortality_paper

## Notes

### Competing Interest Statement

The authors have declared no competing interest.

### Funding Statement

This study was funded by the Dutch public funding agency ZonMw, grant number 10430252220010.

### Author Declarations

The IRB of University Medical Centre Utrecht waived ethical approval for this work. This study does not fall under the scope of the Dutch Medical Research Involving Human Subjects Act (WMO). It therefore does not require approval from an accredited medical ethics committee in the Netherlands. However, in the UMC Utrecht, an independent quality check has been carried out to ensure compliance with legislation and regulations (regarding Informed Consent procedure, data management, privacy aspects and legal aspects).

