## Supplemental for "COVID-19 vaccination and short-term mortality risk: a nationwide self-controlled case series study in The Netherlands"

### Supplemental file

### Supplemental tables

**Supplemental table 1.** Characteristics of individuals in the general Dutch population died from any cause between 6 January 2021 and 18 November 2021, stratified by risk period in which death occurred after last COVID-19 vaccination dose received.

| **Characteristic** | **Timing of death** | | |
|---|---|---|---|
| | **Risk period after COVID-19 vaccination** | | **No registered COVID-19 vaccination** |
| | **≤3 weeks** | **>4 weeks** | |
| **Total** | 8,073 | 67,348 | 66,291 |
| Mean age (SD), years | 79.15 (12.17) | 79.84 (11.72) | 75.73 (14.75) |
| Age group | | | |
| <29 | 32 (0.4) | 202 (0.3) | 857 (1.3) |
| 30-39 | 41 (0.5) | 236 (0.4) | 770 (1.2) |
| 40-49 | 117 (1.4) | 745 (1.1) | 1,719 (2.6) |
| 50-59 | 355 (4.4) | 2790 (4.1) | 5,036 (7.6) |
| 60-69 | 956 (11.8) | 7453 (11.1) | 9,912 (15.0) |
| 70-79 | 2,077 (25.7) | 16,392 (24.3) | 17,576 (26.5) |
| 80-89 | 2,937 (36.4) | 25,996 (38.6) | 20,014 (30.2) |
| ≥90 | 1,558 (19.3) | 13,534 (20.1) | 10,407 (15.7) |
| Women, n (%) | 4,022 (49.8) | 34,590 (51.4) | 32,153 (48.5) |
| Cause of death | | | |
| Circulatory system diseases | 2,341 (29.0) | 16,158 (24.0) | 13,275 (20.0) |
| Neoplasms | 1,536 (19.0) | 19,127 (28.4) | 20,002 (30.2) |
| Respiratory system diseases | 510 (6.3) | 4,473 (6.6) | 3,488 (5.3) |
| Mental and neurological diseases | 1,087 (13.5) | 10,313 (15.3) | 5,578 (8.4) |
| Non-natural causes | 541 (6.7) | 3,781 (5.6) | 3,517 (5.3) |
| Other causes | 1,308 (16.2) | 10,860 (16.1) | 10,442 (15.8) |
| COVID-19 | 750 (9.3) | 2,636 (3.9) | 9,989 (15.1) |
| Number of vaccine administrations | | | |
| One dose | 4,395 (54.4) | 11,317 (16.8) | |
| Two doses | 3,565 (44.2) | 55,927 (83.0) | |
| Three doses | 113 (1.4) | 104 (0.2) | |
| Vaccine type, first vaccine | | | |
| mRNA | 7,250 (89.8) | 61,123 (90.8) | |
| non-mRNA or unknown | 823 (10.2) | 6,222 (9.2) | |
| Vaccine type, second vaccine | | | |
| mRNA | 3,283 (40.7) | 52,260 (77.6) | |
| non-mRNA or unknown | 396 (4.9) | 3,772 (5.6) | |
| Vaccine type, third vaccine | | | |
| mRNA | 109 (1.4) | 55 (0.1) | |
| non-mRNA or unknown[1] | 5 (0.0) | 50 (0.1) | |
| Presence of chronic conditions | | | |
| No | 1,754 (21.7) | 15,422 (22.9) | 17,233 (26.0) |
| One | 1,440 (17.8) | 13,355 (19.8) | 12,828 (19.4) |
| Multiple | 4,879 (60.4) | 38,571 (57.3) | 36,230 (54.7) |
| Chronic condition | | | |
| Anaemia | 1,744 (21.6) | 12,979 (19.3) | 12,510 (18.9) |
| Asplenia[2] | 40 (0.1) | | 37 (0.1) |
| Asthma | 277 (3.4) | 2,025 (3.0) | 2,024 (3.1) |
| Chronic Liver Disease | 153 (1.9) | 1,080 (1.6) | 1,367 (2.1) |
| Cardiovascular Disease | 3,934 (48.7) | 31,155 (46.3) | 28,398 (42.8) |
| Diabetes | 1,591 (19.7) | 11,926 (17.7) | 11,713 (17.7) |
| Hypertension | 3,045 (37.7) | 23,700 (35.2) | 21,651 (32.7) |

| | | | |
|---|---|---|---|
| Obesity | 261 (3.2) | 1,910 (2.8) | 2,043 (3.1) |
| Immunodeficiency and organ transplant | 92 (1.1) | 847 (1.3) | 820 (1.2) |
| Neuromuscular Disorders | 13 (0.2) | 70 (0.1) | 54 (0.1) |
| Renal diseases | 1,689 (20.9) | 12,548 (18.6) | 12,120 (18.3) |
| Dementia | 1,477 (18.3) | 9,988 (14.8) | 7,617 (11.5) |
| Stroke | 414 (5.1) | 2,969 (4.4) | 2,746 (4.1) |
| Rheumatic disorders | 237 (2.9) | 1,697 (2.5) | 1,598 (2.4) |
| Cancer | 2,161 (26.8) | 19,584 (29.1) | 19,934 (30.1) |
| Lung disease (excluding asthma) | 1,856 (23.0) | 14,132 (21.0) | 14,291 (21.6) |
| Tuberculosis[2] | 24 (0.0) | | 39 (0.1) |
| Number of registered positive SARS-CoV-2 infections | | | |
| 0 | 7,106 (88.0) | 60,894 (90.4) | 54,877 (82.8) |
| 1 | 961 (11.9) | 6,383 (9.5) | 11,366 (17.1) |
| 2 or 3 | <10 | >67 | 48 (0.1) |
| Type of COVID-19 test, first test[3] | | | |
| PCR | >956 | > 6,430 | 11,389 (99.9) |
| Other | <10 | < 20 | 14 (0.1) |

[1] Rounded to nearest fifth due to low counts.
[2] Presence of chronic conditions shown for two categories combined due to low counts.
[3]All second and third tests were PCR.

**Supplemental table 2.** Characteristics of individuals in the general Dutch population who received at least one COVID-19 vaccination and died from any cause between 6 January 2021 and 30 April 2023.

| **Characteristic** | **Vaccinated** | | | **No vaccine registered** |
|---|---|---|---|---|
| | **Total** | **Risk period after vaccination** | | |
| | | **≤3 weeks** | **>4 weeks** | |
| **Total** | 299,935 | 17,991 | 281,944 | 96,830 |
| Mean age (SD), years | 78.35 (12.27) | 79.95 (11.51) | 78.25 (12.31) | 74.78 (15.75) |
| Age group | | | | |
| <29 | 1,341 (0.4) | 49 (0.3) | 1,293 (0.5) | 1,774 (1.8) |
| 30-39 | 1,554 (0.5) | 68 (0.4) | 1,486 (0.5) | 1,510 (1.6) |
| 40-49 | 4,494 (1.5) | 194 (1.1) | 4,300 (1.5) | 2,986 (3.1) |
| 50-59 | 15,321 (5.1) | 625 (3.5) | 14,696 (5.2) | 8,003 (8.3) |
| 60-69 | 37,511 (12.5) | 1,960 (10.9) | 35,551 (12.6) | 14,429 (14.9) |
| 70-79 | 79,573 (26.5) | 4,583 (25.5) | 74,990 (26.6) | 24,667 (25.5) |
| 80-89 | 110,156 (36.7) | 6,927 (38.5) | 103,229 (36.6) | 29,023 (30.0) |
| ≥90 | 49,985 (16.7) | 3,583 (19.9) | 46,402 (16.5) | 14,438 (14.9) |
| Women, n (%) | 152,512 (50.8) | 9,264 (51.5) | 143,248 (50.8) | 47,332 (48.9) |
| Cause of death | | | | |
| Circulatory system diseases | 70,305 (23.4) | 5,297 (29.4) | 65,008 (23.1) | 19,364 (20.0) |
| Neoplasms | 82,379 (27.5) | 3,050 (17.0) | 79,329 (28.1) | 26,930 (27.8) |
| Respiratory system diseases | 23,118 (7.7) | 1,406 (7.8) | 21,712 (7.7) | 5,490 (5.7) |
| Mental and neurological diseases | 41,235 (13.7) | 2,636 (14.7) | 38,599 (13.7) | 8,454 (8.7) |
| Non-natural causes | 17,557 (5.9) | 1,209 (6.7) | 1,6348 (5.8) | 5,790 (6.0) |
| Other causes | 50,616 (16.9) | 3,044 (16.9) | 47,572 (16.9) | 16,806 (17.4) |
| COVID-19 | 14,725 (4.9) | 1,349 (7.5) | 13,376 (4.7) | 13,996 (14.5) |
| Number of vaccinations | | | | |
| One dose | 22,682 (7.6) | 4,659 (25.9) | 18,023 (6.4) | |
| Two doses | 102,043 (34.0) | 3,886 (21.6) | 98,157 (34.8) | |
| Three doses | 82,166 (27.4) | 4,533 (25.2) | 77,633 (27.5) | |
| Four doses | 57,586 (19.2) | 2,928 (16.3) | 54,658 (19.4) | |
| Five doses | 34,178 (11.4) | 1,918 (10.7) | 32,260 (11.4) | |
| Six or seven doses | 1,280 (0.4) | 67 (0.4) | 1,213 (0.4) | |
| Completed primary series | 275,559 (91.9) | 275,559 (73.7) | 275,559 (93.0) | |
| Number of booster events[1] | | | | |
| None | 213,467 (53.8) | 8,608 (47.8) | 108,029 (38.3) | |
| One | 86,496 (21.8) | 4,042 (22.5) | 82,454 (29.2) | |
| Two | 58,874 (14.8) | 3,206 (17.8) | 55,668 (19.7) | |
| Three | 35,391 (8.9) | 1,979 (11.0) | 33,412 (11.9) | |
| Four or five | 2,537 (0.6) | 156 (0.9) | 2,371 (0.8) | |
| Vaccin type, first vaccine | | | | |
| mRNA | 266,482 (88.8) | 16,060 (89.3) | 250,422 (88.8) | |
| non-mRNA or unknown | 28,808 (9.6) | 1,613 (9.0) | 27,195 (9.6) | |
| Vaccin type, second vaccine | | | | |
| mRNA | 247,618 (82.6) | 11,805 (65.6) | 235,813 (83.6) | |
| non-mRNA or unknown | 22,391 (7.5) | 1,088 (6.0) | 21,303 (7.6) | |
| Vaccin type, third vaccine | | | | |
| mRNA | 20,722 (6.9) | 991 (5.5) | 19,731 (7.0) | |
| non-mRNA or unknown | 307 (0.1) | 15 (0.1) | 292 (0.1) | |
| Vaccin type, first booster | | | | |
| mRNA | 17,272 (57.6) | 9,432 (52.4) | 16,329 (57.9) | |
| non-mRNA or unknown[2] | 5 (0.0) | 0 (0.0) | 5 (0.0) | |
| Vaccin type, second booster | | | | |
| mRNA | 89,862 (30.0) | 4,836 (26.9) | 85,026 (30.2) | |

| | | | | |
|---|---|---|---|---|
| non-mRNA or unknown[2] | 5 (0.0) | 0 (0.0) | 5 (0.0) | |
| Vaccin type, third booster | | | | |
| mRNA | 33,060 (11.0) | 1,881 (10.5) | 31,179 (11.1) | |
| non-mRNA or unknown[2] | 5 (0.0) | 0 (0.0) | 5 (0.0) | |
| Vaccin type, fourth booster | | | | |
| mRNA | 183 (0.1) | 12 (0.1) | 171 (0.1) | |
| non-mRNA or unknown | 0 (0.0) | 0 (0.0) | 0 (0.0) | |
| Number of registered COVID-19 infections | | | | |
| 0 | 265,807 (88.6) | 15,719 (87.4) | 250,088 (88.7) | 80,996 (83.6) |
| 1 | 33,720 (11.2) | 2,242 (12.5) | 31,478 (11.2) | 15,723 (16.2) |
| 2 or 3 | 408 (0.1) | 30 (0.2) | 378 (0.1) | 111 (0.1) |

[1] Booster events were defined as a booster vaccination or a registered positive SARS-CoV-2 after completion of the primary series vaccination.
[2] Rounded to nearest fifth due to low counts.

**Supplemental table 3.** Characteristics of individuals in the general Dutch who died from any cause between 1 June 2020 and 31 December 2021, stratified by risk period in which death occurred after a registered positive SARS-CoV-2 infection.

| Characteristic | Timing of death | | |
|---|---|---|---|
| | Risk period after registered positive SARS-CoV-2 infection | | No registered SARS-CoV-2 infection |
| | ≤3 weeks | >4 weeks | |
| **Total** | 21,243 | 12,632 | 229,706 |
| Mean age (SD), years | 80.8 (10.2) | 77.5 (13.5) | 77.5 (13.5) |
| Age group, n (%) | | | |
| <29 | 36 (0.2) | 115 (0.9) | 1,483 (0.8) |
| 30-39 | 43 (0.2) | 77 (0.6) | 1,859 (0.8) |
| 40-49 | 142 (0.7) | 235 (1.9) | 4,588 (2.0) |
| 50-59 | 540 (2.5) | 794 (6.3) | 13,844 (6.0) |
| 60-69 | 1,834 (8.6) | 1,744 (13.8) | 30,647 (13.3) |
| 70-79 | 5,655 (26.6) | 3,181 (25.2) | 58,885 (25.6) |
| 80-89 | 9,084 (42.8) | 4,247 (33.6) | 79,232 (34.5) |
| ≥90 | 3,909 (18.4) | 2,241 (17.7) | 38,821 (16.9) |
| Women, n (%) | 9,628 (45.3) | 6518 (51.6) | 115370 (50.2) |
| Cause of death | | | |
| Circulatory system diseases | 503 (2.4) | 2,316 (18.3) | 55,040 (24.0) |
| Neoplasms | 370 (1.7) | 2,294 (18.2) | 71,747 (31.2) |
| Respiratory system diseases | 111 (0.5) | 558 (4.4) | 14,339 (6.2) |
| Mental and neurological diseases | 214 (1.0) | 1,703 (13.5) | 29,091 (12.7) |
| Non-natural causes | 266 (1.3) | 691 (5.5) | 13,496 (5.9) |
| Other causes | 738 (3.5) | 1,775 (14.1) | 38,517 (16.8) |
| COVID-19 | 19,041 (89.6) | 3,295 (26.1) | 7,476 (3.3) |
| Number of registered positive SARS-CoV-2 infections | | | |
| 1 | 21,139 (99.5) | 12,546 (99.3) | |
| 2 or 3 | 104 (0.5) | 85 (0.7) | |
| Type of COVID-19 test, first test[1,2] | | | |
| PCR | 21,186 (99.8) | 12,585 (99.7) | |
| Self-test | 22 (0.1) | 21 (0.2) | |
| Presence of chronic conditions | | | |
| No | 6,063 (28.5) | 4,152 (32.9) | 79,804 (34.7) |
| One | 3,965 (18.7) | 2,729 (21.6) | 53,593 (23.3) |
| Multiple | 11,215 (52.8) | 5,751 (45.5) | 96,309 (41.9) |
| Chronic condition | | | |
| Anaemia | 4,441 (20.9) | 2,304 (18.2) | 38,053 (16.6) |
| Asplenia[3] | 18 (0.1) | | 90 (0.0) |
| Asthma | 823 (3.9) | 460 (3.6) | 5,792 (2.5) |
| Chronic Liver Disease | 286 (1.3) | 194 (1.5) | 3,752 (1.6) |
| Cardiovascular Disease | 10,863 (51.1) | 5,705 (45.2) | 96,744 (42.1) |
| Diabetes | 5,003 (23.6) | 2,444 (19.3) | 37,251 (16.2) |
| Hypertension | 8,316 (39.1) | 4,444 (35.2) | 71,119 (31.0) |
| Obesity | 828 (3.9) | 421 (3.3) | 5,620 (2.4) |
| Immunodeficiency and organ transplant | 318 (1.5) | 181 (1.4) | 2,468 (1.1) |
| Neuromuscular Disorders | 48 (0.2) | 15 (0.1) | 192 (0.1) |
| Renal diseases | 4,868 (22.9) | 2,334 (18.5) | 37,733 (16.4) |
| Dementia | 4,243 (20.0) | 2,063 (16.3) | 26,865 (11.7) |
| Stroke | 1,219 (5.7) | 591 (4.7) | 8,959 (3.9) |
| Rheumatic disorders | 717 (3.4) | 332 (2.6) | 5,176 (2.3) |
| Cancer | 4,348 (20.5) | 3,132 (24.8) | 61,542 (26.8) |

| | | | |
|---|---|---|---|
| Lung disease (excluding asthma) | 4,967 (23.4) | 2,543 (20.1) | 43,867 (19.1) |
| Tuberculosis[3] | 22 (0.1) | | 92 (0.0) |
| Vaccine type, first vaccine | | | |
| mRNA | 5,634 (26.5) | 5,973 (47.3) | 75,209 (32.7) |
| non-mRNA or unknown | 329 (1.5) | 458 (3.6) | 8,219 (3.6) |
| Vaccine type, second vaccine | | | |
| mRNA | 4,514 (21.2) | 4,330 (34.3) | 64,438 (28.1) |
| non-mRNA or unknown | 263 (1.2) | 221 (1.7) | 5,335 (2.3) |
| Vaccine type, third vaccine | | | |
| mRNA | 129 (0.6) | 65 (0.5) | 1227 (0.5) |
| non-mRNA or unknown[4] | 5 (0.0) | 5 (0.0) | 70 (0.0) |

[1] Antigen and IgM tests not shown due to low counts.
[2] All second and third tests were PCR.
[3] Presence of chronic conditions shown for two categories combined due to low counts.
[4] Rounded to nearest fifth due to low counts.

**Supplemental table 4.** Sensitivity analyses of relative incidence of deaths in the 3-week risk period following basic series COVID-19 vaccination, all doses combined, or a registered positive SARS-CoV-2 infection

| | **All-cause deaths**<br>Relative incidence (95%CI) | **Non-COVID-19 related deaths**<br>Relative incidence (95%CI) |
|---|---|---|
| **Sensitivity analyses for vaccination** | | |
| **Exclusion individuals with a registered positive SARS-CoV-2 infection** in the 8 weeks before vaccination or during the exposure period (*n* = 2,662) | 0.52 (0.51-0.54) | 0.54 (0.53-0.56) |
| **Calendar time adjustment** with a restricted cubic spline | 0.46 (0.45-0.47) | 0.45 (0.44-0.47) |
| **No adjustment for calendar time** | 0.59 (0.57-0.60) | 0.55 (0.54-0.56) |
| **Risk period** | | |
| Week 1-12 | 0.71 (0.70-0.72) | 0.71 (0.69-0.72) |
| Week 1 | 0.29 (0.27-0.30) | 0.30 (0.28-0.32) |
| Week 2 | 0.49 (0.47-0.51) | 0.48 (0.46-0.50) |
| Week 3 | 0.63 (0.61-0.65) | 0.59 (0.57-0.61) |
| Week 4 | 0.67 (0.64-0.69) | 0.64 (0.62-0.67) |
| Week 5 | 0.75 (0.72-0.78) | 0.73 (0.70-0.76) |
| Week 6 | 0.92 (0.89-0.96) | 0.91 (0.88-0.95) |
| Week 7 | 0.91 (0.88-0.95) | 0.90 (0.87-0.94) |
| Week 8 | 0.85 (0.81-0.88) | 0.85 (0.82-0.89) |
| Week 9 | 0.86 (0.83-0.90) | 0.87 (0.84-0.91) |
| Week 10 | 0.81 (0.78-0.85) | 0.82 (0.79-0.86) |
| Week 11 | 0.83 (0.80-0.87) | 0.84 (0.80-0.87) |
| Week 12 | 0.81 (0.78-0.85) | 0.82 (0.78-0.85) |
| **Administrator** | | |
| Municipal health service | 0.46 (0.44-0.48) | 0.45 (0.43-0.47) |
| General practitioner | 0.55 (0.52-0.59) | 0.56 (0.52-0.60) |
| Other (nursing homes, residential care centres, hospitals, mobile units) | 0.77 (0.74-0.80) | 0.78 (0.75-0.81) |
| **Sensitivity analyses for infection** | | |
| **Calendar time adjustment** with a restricted cubic spline | 13.18 (12.85-13.53) | 1.77 (1.68-1.87) |
| **No adjustment for calendar time** | 17.10 (16.68-17.53) | 2.10 (1.99-2.22) |
| **Risk period** | | |
| Week 1-12 | 8.28 (8.05-8.51) | 1.54 (1.47-1.60) |
| Week 1 | 25.54 (24.68-26.44) | 2.63 (2.43-2.85) |
| Week 2 | 34.15 (33.05-35.28) | 2.57 (2.37-2.78) |
| Week 3 | 15.77 (15.16-16.40) | 2.01 (1.84-2.20) |
| Week 4 | 7.42 (7.05-7.81) | 1.57 (1.42-1.74) |
| Week 5 | 4.75 (4.46-5.06) | 1.61 (1.45-1.78) |
| Week 6 | 3.22 (2.99-3.47) | 1.28 (1.13-1.43) |
| Week 7 | 2.47 (2.26-2.69) | 1.36 (1.21-1.53) |
| Week 8 | 1.86 (1.68-2.05) | 1.18 (1.03-1.34) |
| Week 9 | 1.55 (1.39-1.74) | 1.07 (0.94-1.23) |
| Week 10 | 1.49 (1.32-1.67) | 1.22 (1.07-1.39) |
| Week 11 | 1.20 (1.05-1.37) | 0.98 (0.84-1.13) |
| Week 12 | 1.19 (1.05-1.35) | 1.04 (0.91-1.19) |
| **Induction interval** | | |
| 2 days before registration date | 14.79 (14.42-15.17) | 2.05 (1.94-2.16) |
| 4 days before registration date | 13.02 (12.70-13.36) | 1.86 (1.76-1.97) |
| 7 days before registration date | 9.91 (9.66-10.17) | 1.59 (1.50-1.69) |
| 14 days before registration date | 2.54 (2.46-2.62) | 0.82 (0.76-0.89) |

### Supplemental figures

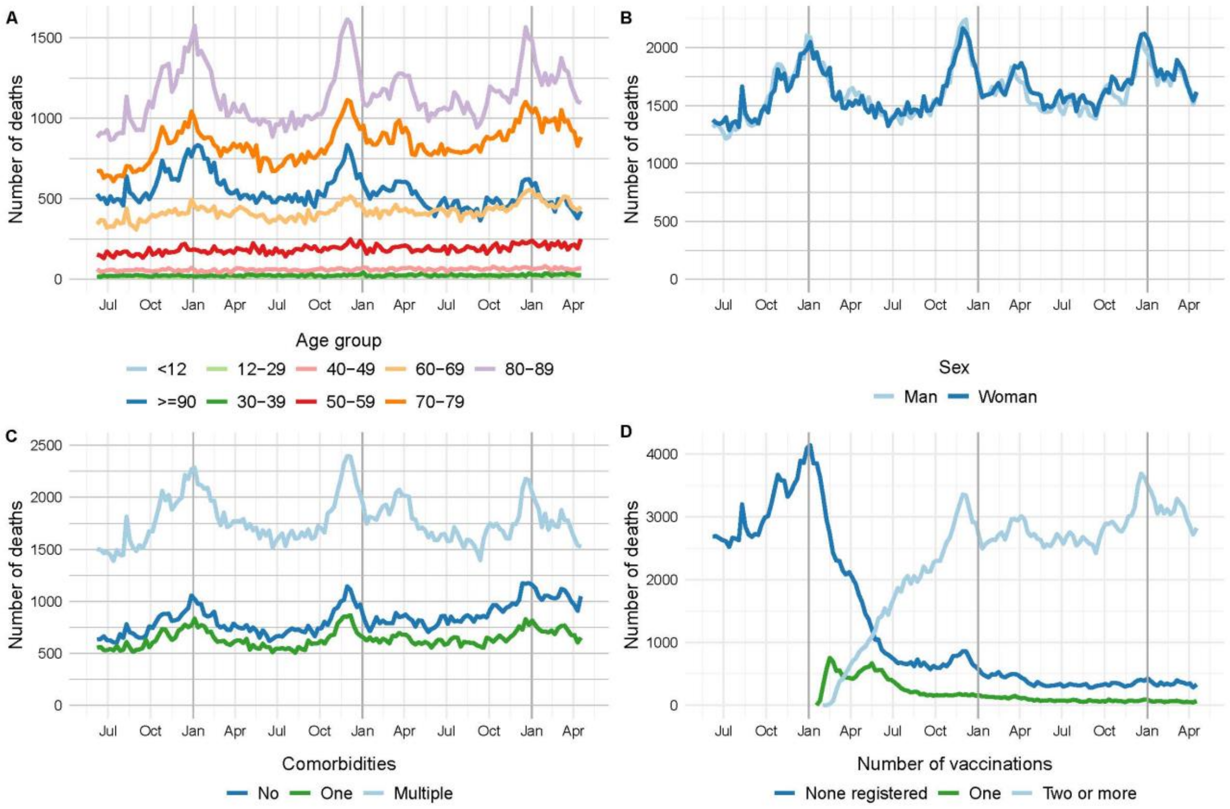


**Supplemental figure 1.** Number of deaths each week, among individuals who died between June 1, 2020 to April 30, 2023 (*n* = 493,382) by, A) by age group, B) sex, C) presence of chronic disease, and D) vaccination doses received.

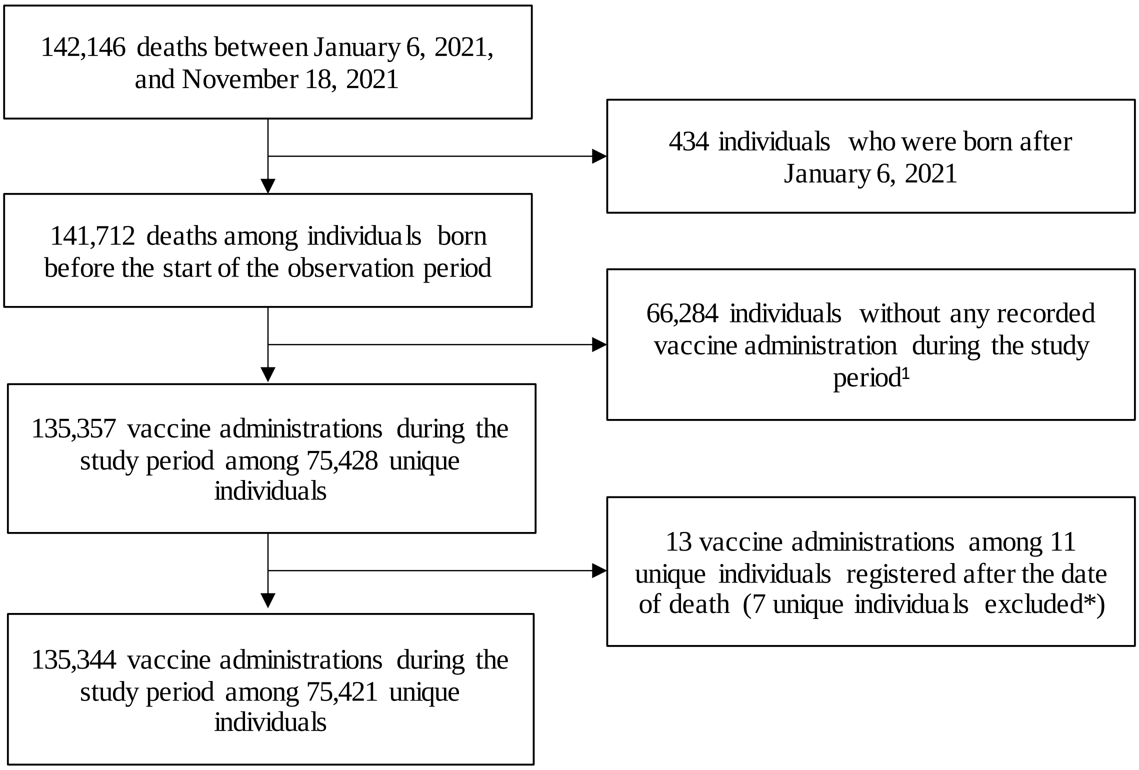


**Supplemental figure 2.** Flow-chart of the study population for analysis on COVID-19 vaccination and mortality.

[1] Included in the analyses to control for seasonal trends in mortality.

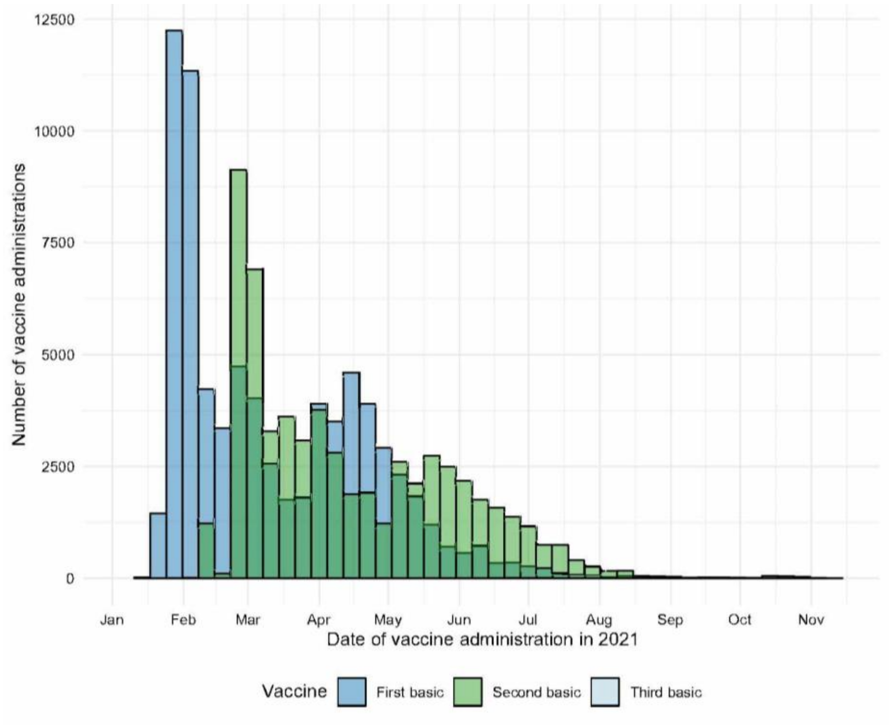


**Supplemental figure 3a.** Number of primary vaccine administrations from January 6, 2021 to November 18, 2021, among individuals who received the vaccine and died during this period (*n* = 75,421).

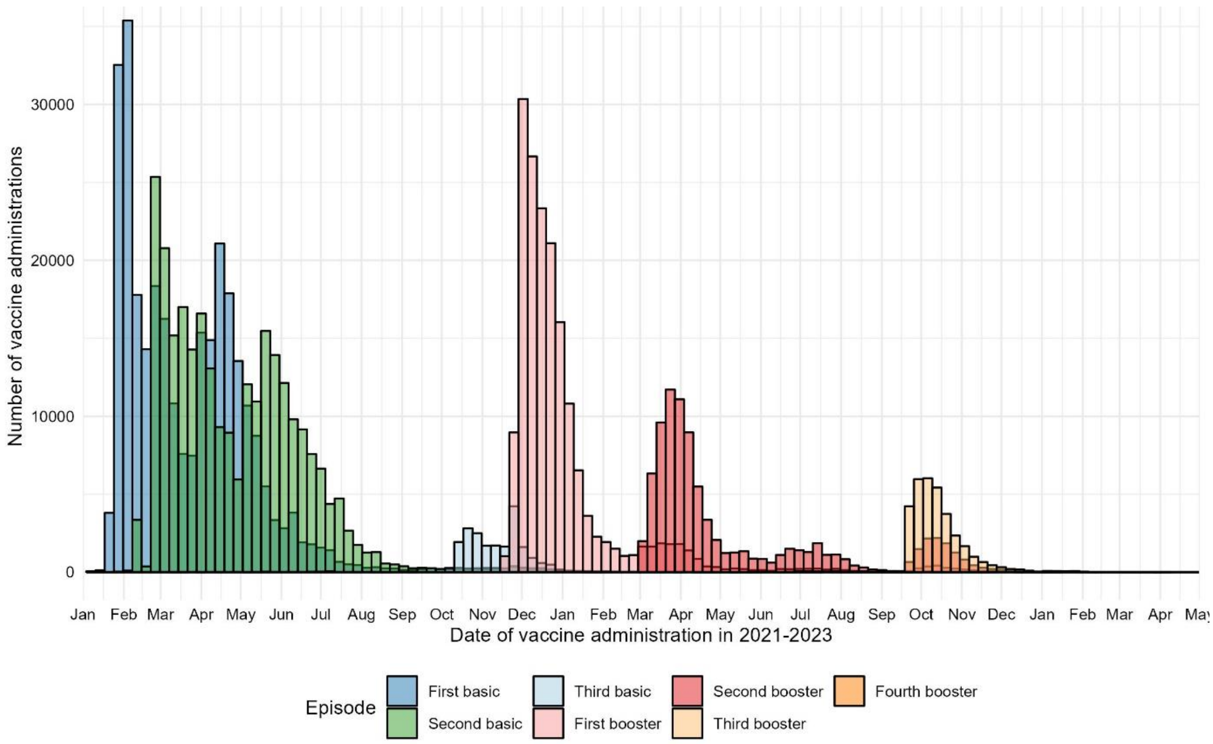


**Supplemental figure 3b.** Number of primary and booster vaccine administrations from January 6, 2021 to April 30, 2023, among individuals who received the vaccine and died during this period (*n* = 299,935).

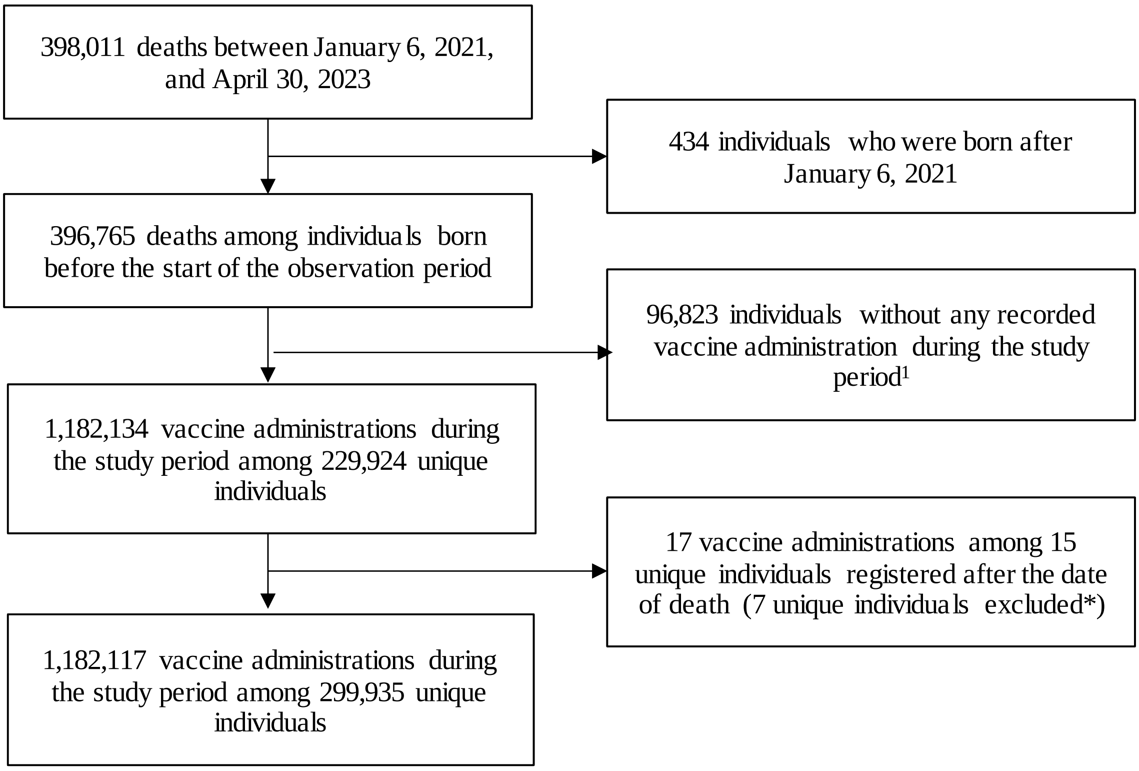


**Supplemental figure 4.** Flow-chart of the study population for analysis on COVID-19 basic and booster vaccination and mortality.

[1] Included in the analyses to control for seasonal trends in mortality.

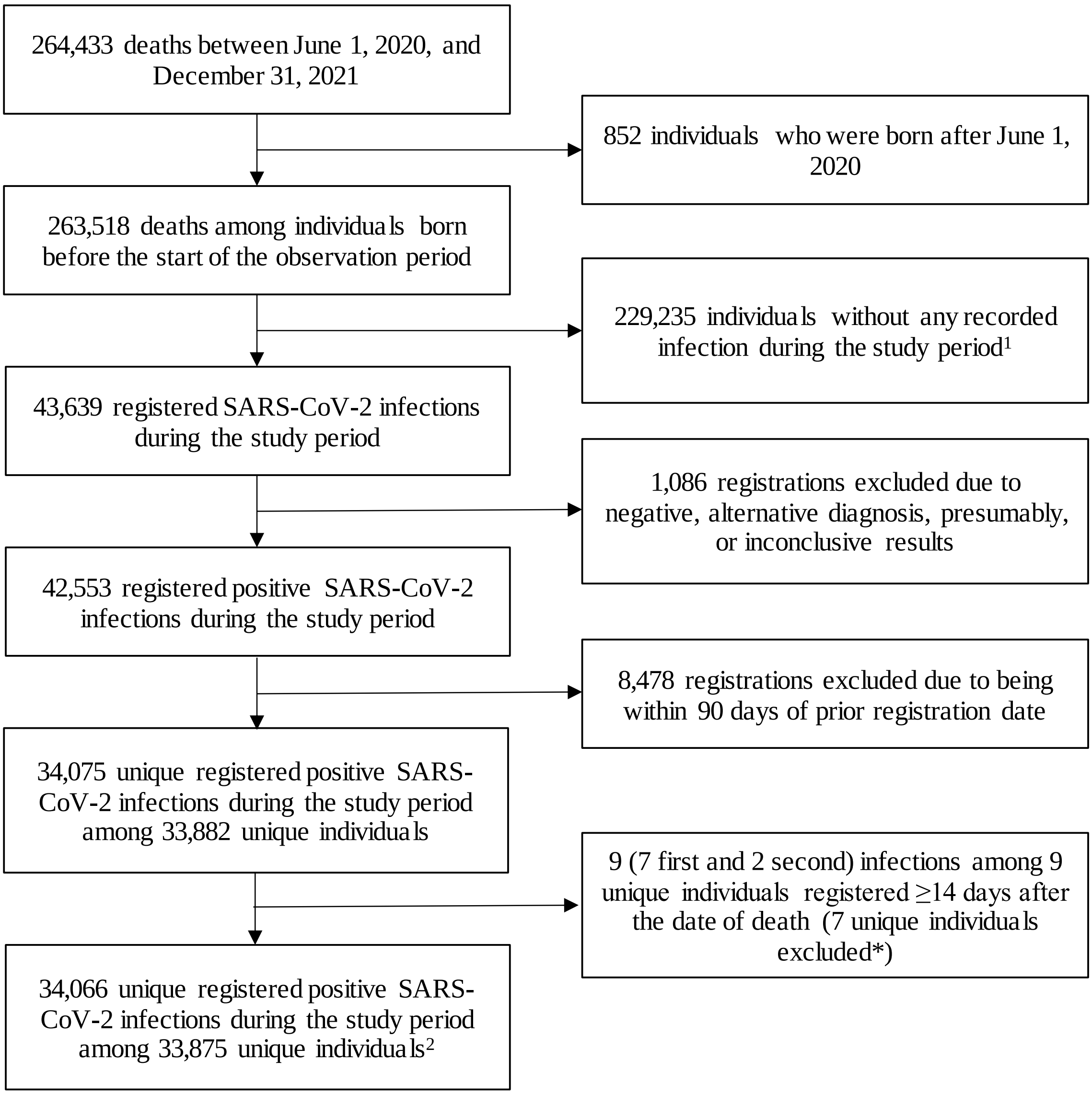


**Supplemental figure 5.** Flow-chart of the study population for analysis on SARS-CoV2 infection and mortality.

[1] Included in the analyses to control for seasonal trends in mortality.

[2] Including 240 (236 first and 4 second) infections among 240 unique individuals within 14 days after the date of death.

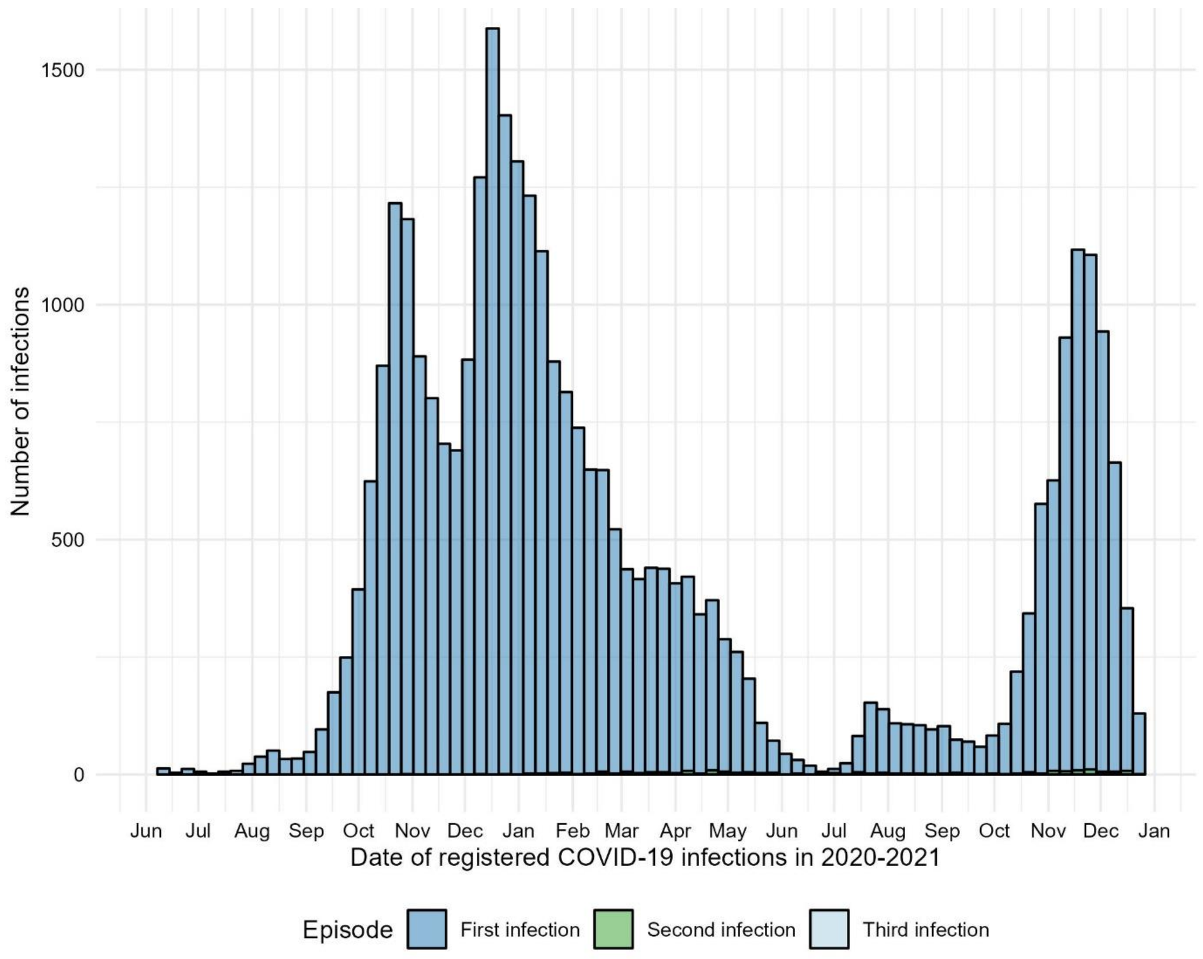


**Supplemental figure 6.** Number of registered positive SARS-CoV-2 infections from June 1, 2020 to December 31, 2021, among individuals who had a registered positive SARS-CoV-2 infection and died during this period ($n$ = 33,875).
